# Microsomal reductase activity in patients with thyroid neoplasms

**DOI:** 10.1101/2020.06.09.20126672

**Authors:** Elena V. Proskurnina, Maria V. Fedorova, Madina M. Sozarukova, Aleksandr E. Mitichkin, Igor V. Panteleev, Evgeny V. Svetlov

**Affiliations:** Research Centre for Medical Genetics, Moscow, Russia; Faculty of Fundamental Medicine, Lomonosov Moscow State University, Moscow, Russia; Kurnakov Institute of General and Inorganic Chemistry of the Russian Academy of Sciences, Moscow, Russia; Inozemtsev City Clinical Hospital of Moscow Department of Health, Moscow, Russia

**Keywords:** Papillary thyroid cancer, cytochrome b5 reductase, cytochrome P450 reductase, chemiluminescence

## Abstract

**Objective:** Cytochrome b5 reductase (CYB5R) and cytochrome P450 reductase (CYPOR) play an important role in cell metabolism; however, their role in thyroid hormonogenesis and carcinogenesis has not been studied. The activity of CYB5R correlates with metastasizing in breast cancer, but there are no similar studies for CYB5R and CYPOR for thyroid cancer. The aim was to assess the activity of CYB5R and CYPOR in thyroid tissues in benign and malignant thyroid neoplasms.

**Methods:** 36 patients with thyroid neoplasms participated in the study. The control euthyroid goiter group included 10 patients; the thyrotoxic nodular goiter group included 14 patients; the papillary thyroid cancer T1-2N0-1M0 (PTC) group included 12 patients. The activity of CYB5R and CYPOR was assessed with lucigenin-enhanced chemiluminescence stimulated by NADH and NADPH, respectively.

**Results:** The activity of CYB5R and CYPOR increased several times in thyrotoxicosis and approximately by an order of magnitude in some cases of PTC, but the activity change of CYPOR was more pronounced compared to CYB5R. For the PTC group, the subgroups with low and high activity of microsomal reductases were detected. Microsomal reductases in follicular adenoma was 2–4-fold less active compared to nontoxic goiter and the low-activity PTC group.

**Conclusions:** Activity of microsomal reductases varies in thyroid pathology and can serve as a diagnostic and prognostic parameter in papillary thyroid cancer.

## Introduction

Thyroid neoplasms are detected in 20–50% of cases of thyroid pathology, 4–5% of cases of which are cancer. Although thyroid cancer is quite rare and makes up from 1 to 4% of all malignant tumors, morbidity of thyroid cancer has increased 2.5 times recently [1]. The development of new approaches to diagnosing and predicting the development of this type of cancer is of special interest, and the search for early markers of metastasizing is one of the priority tasks of oncology.

The carcinogenesis in the thyroid gland tissue is a complex process which was not fully understood. However, it is clear that reactive oxygen species (ROS) plays important and ambiguous role [2-4]. It is due to the participation of hydrogen peroxide in the synthesis of thyroid hormones. On the one hand, ROS can activate carcinogenic signaling pathways, promote the survival and proliferation of cancer cells, lead to DNA damage and genome instability [5]. On the other hand, ROS activate antitumor signaling pathways and provide programmed cell death [6] [7]. Switching between the pro- and anti-carcinogenic effects of ROS depends, inter alia, on their concentration.

Oxidative stress in thyroid cancer and thyrotoxicosis is demonstrated by an increased level of malondialdehyde that is lipid peroxidation marker [4] [2]. In hyperthyroidism, this is the result of hyperproduction of thyroid hormones, and in cancer oxidative stress is caused by an active proliferation of malignant cells for which ROS are invasion and transformation factors. Microsomal respiratory chains are one of the most prominent sources of intracellular superoxide anion radical and hydrogen peroxide. The final protein of the NADPH-dependent microsomal chain is cytochrome P450 (CYP); the final protein of the NADH-dependent microsomal chain is cytochrome b5 (CYB5). Today, NADH-dependent cytochrome b5 reductase (CYB5R) is of special interest. This enzyme is involved in the synthesis of cholesterol, elongation of fatty acids, microsomal hydroxylation of xenobiotics and steroid hormones. It is also a part of the transmembrane redox system that provides reduced ascorbate and coenzyme Q10 and protects the cell from apoptosis [8]. Besides reducing CYP, NADPH-dependent cytochrome P450 reductase (CYPOR) can transfer an electron to cytochrome b5, heme oxygenase, squalene-monooxygenase, and 7-dehydrocholesterol reductase. NADPH-dependent microsomal chain provides the metabolism of prodrugs, especially anticancer drugs [9].

The role of CYB5R in carcinogenesis has not been studied well. The increased expression of CYB5R is known to correlate with poor prognosis in patients with estrogen receptor-negative breast cancer; a decrease in CYB5R gene expression significantly reduced lung metastasis in mice [10]. As for CYPOR, the available work is mainly devoted to the participation of this enzyme in the metabolism of anticancer drugs [11] [12]. We think that the information on the activity of CYB5R and CYPOR can be useful for understanding the mechanisms of hormonogenesis and carcinogenesis. It will also help in solving diagnostic and prognostic tasks. Lucigenin-enhanced chemiluminescence stimulated by NADH and NADPH is a promising method for studying the activity of CYB5R and CYPOR, respectively. Lucigenin is directly reduced by these enzymes. In the presence of oxygen, a superoxide radical anion is formed. The intensity of chemiluminescence should be proportional to the activity of CYB5R and CYPOR [13,14] [15].

## Methods

### Participants

36 patients with thyroid neoplasms (mean age 50.3 ± 12.7 y.o.) participated in the study. The control euthyroid goiter group included 10 patients (mean age 53.3 ± 8.4 y.o.); the thyrotoxic nodular goiter group included 14 patients (mean age 47.3 ± 13.4 y.o.), the papillary thyroid cancer T1-2N0-1M0 (PTC) group included 12 patients (mean age 51.7 ± 14.1 y.o.). Among the of benign cases, there were *n* = 2 cases of Hürthle cell adenoma (toxic goiter) and *n* = 4 cases of follicular adenoma (euthyroid goiter). Before surgery treatment, the patients with thyroid cancer did not receive radioiodine or other therapy. There were patients with autoimmune thyroiditis *n* = 2 in the euthyroid goiter group, *n* = 4 in the PTC group.

Free thyroxine serum levels (T4), thyroid-stimulating hormone (TSH), and thyroid peroxidase antibodies were assessed for every patient. Node size was evaluated by ultrasound sonography with a Toshiba Aplio MX-500 scanner. In patients with nodular pathology, serum calcitonin was quantified, and a fine-needle aspiration biopsy was performed. All the diagnoses were morphologically confirmed.

The patients with confirmed or supposed thyroid cancer and toxic goiter underwent total thyroidectomy; the patients with benign unilateral tumors underwent hemithyroidectomy.

### Assessment of microsomal reductases activity

To assess the activity of tissue cytochrome b5 reductase and cytochrome P450 reductase, we analyzed intra-operative samples of thyroid tissue about 1 × 0.5 cm in size. The samples were kept in a Krebs-Ringer buffer solution at +4°C and analyzed no later than 4 h after taking. To determine the activity of CYB5R and CYPOR, a special protocol was based on the lucigenin-enhanced chemiluminescence and stimulation with NADH or NADPH, respectively. The chemiluminograms were recorded on a Lum-1200 device (DISoft, Russia). Since the chemiluminometer allows to record luminescence in 12 cuvettes, NADH- and NADPH-dependent chemiluminescence were measured in triplicate simultaneously providing high reproducibility and comparability of the results.

The Krebs-Ringer solution (pH 7.4) was prepared daily. As a superoxide-selective chemiluminescence enhancer, lucigenin (10.10-dimethyl-9.9-bacridinium dinitrate, (Sigma, USA) was used. A stock solution (1 mM) was prepared by dissolving a weighed portion in distilled water. Stock solutions of NADH and NADPH (10 mM) (Sigma, USA) were prepared by dissolving weighed portions in distilled water.

Before analysis, the samples were washed three times with Krebs-Ringer solution. Next, equal portions of 15.5 ± 0.5 mg were sampled with a 20G biopsy needle (GTA, Italy). The samples were placed in cuvettes without homogenization. Next, 1860 μL of Krebs-Ringer solution and 120 μL of 1 mM lucigenin were added. Chemiluminescence was recorded at 37°C for 5 min, then 10 μL of 10 mM NADH or NADPH was added, and the signal was recorded for another20 min (Fig. 1). From the chemiluminograms, the basal luminescence intensity *I*_0_, the stimulated luminescence intensity *I*_NADH_ or *I*_NADPH_ were measured. The activation coefficients *K*_NADH_ = (*I*_NADH_ – *I*_0_)/*I*_0_ and *K*_NADPH_ = (*I*_NADPH_–*I*_0_)/*I*_0_ were calculated. The mean of three replicate measurements were calculated for each experiment.

**Figure 1.**
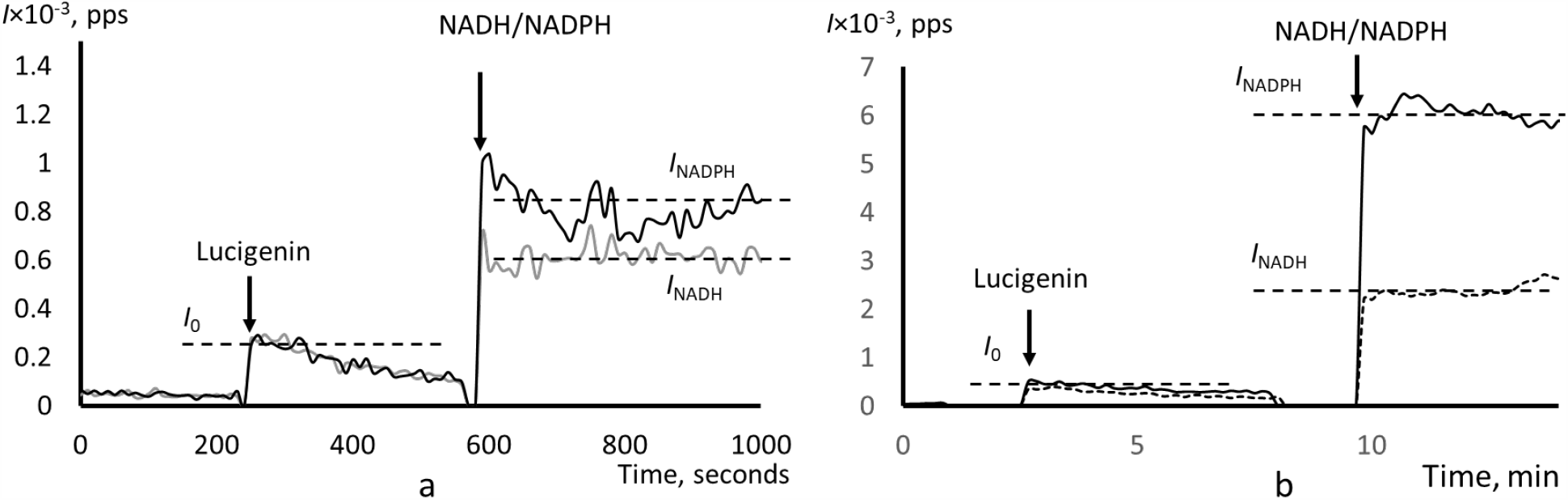
Chemiluminograms of thyroid tissue samples: (a) euthyroid goiter; (b) PTC, a patient with high activity of microsomal reductases; the analytical parameters and time of addition of lucigenin and NADH/NADPH are shown in the figure.

### Statistical Analysis

The size of cohorts was not previously determined. For statistical analysis, STATISTICA for Windows v.10.0 (StatSoft Inc., USA) was used. The normality was checked according to the Shapiro-Wilk test. The values were described as mean 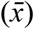 and standard deviation (*S*). Comparative analysis of two independent groups was carried out using the Mann-Whitney test. The relationship was evaluated by the Spearman correlation coefficient (*r*_s_). The differences were considered statistically significant at *p*≤0.05.

## Results

From the chemiluminograms, *I*_NADH_ with *K*_NADH_ = (*I*_NADH_ – *I*_0_)/*I*_0_, and *I*_NADPH_ with *K*_NADPH_ = (*I*_NADPH_ – *I*_0_)/*I*_0_, were determined. These indices represent the activity of CYB5R and CYPOR, respectively.

The *I*_NADH_ and *I*_NADPH_ values for euthyroid and thyrotoxic goiter groups were normally distributed (Shapiro-Wilk test, *p*> 0.20). Patients with autoimmune thyroiditis did not differ from the main group.

In PTC, the distribution functions for all the indicators were asymmetric and bimodal. We classified two subgroups of patients with low (*n* = 6) and high (*n* = 6) activity of CYB5R and CYPOR (Fig. 2). In each subgroup, there were *n* = 2 patients with autoimmune thyroiditis.

**Figure 2.**
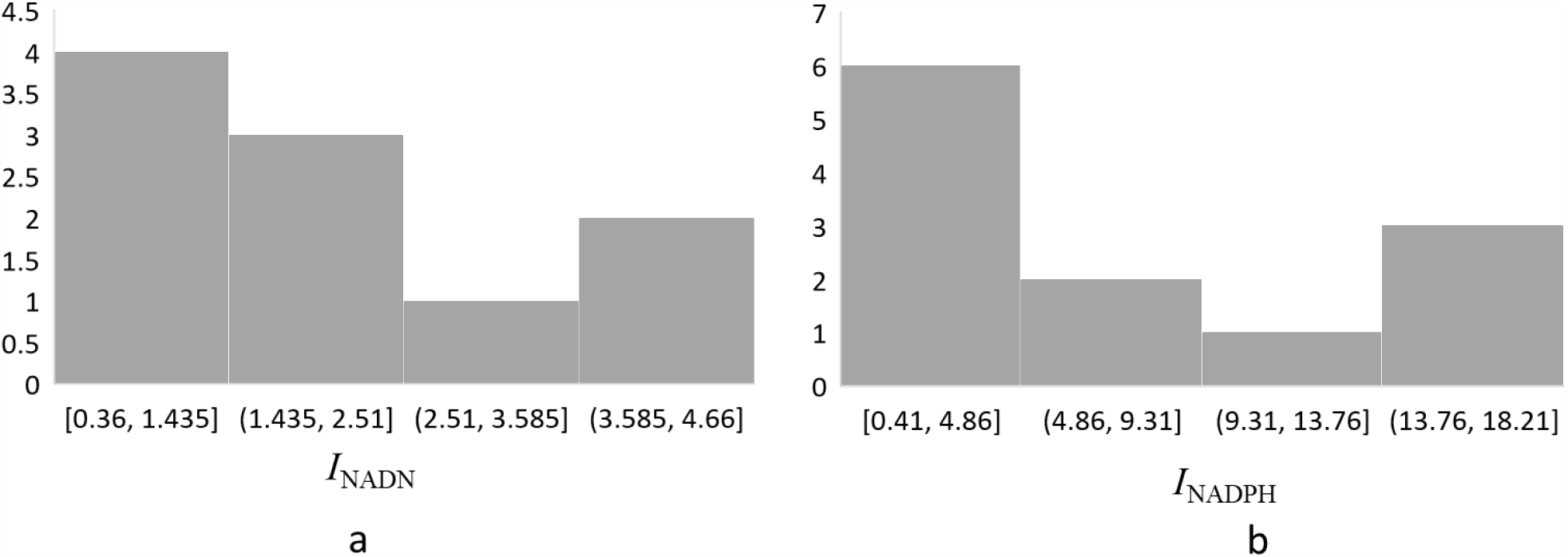
Histograms of (a) *I*_NADH_ and (b) *I*_NADPH_ values for the PTC group; these indices represent the activity of cytochrome b5 reductase and cytochrome P450 reductase, respectively. A comparison of CYB5R and CYPOR activities in euthyroid nodes and neighboring sites did not reveal significant differences (the data not shown). Therefore, we considered the euthyroid-goiter patients as a control group.

Figure 2 demonstrates similar distributions for CYB5R and CYPOR activities in PTC. Moreover, these activities correlated with one another: *r* = 0.72 for *I*_NADH_ *versus I*_NADPH_, *r* = 0.78 for *K*_NADH_ *versus K*_NADPH_. Thus, there were cancer patients with low and high activity of microsomal reductases. To compare, in the groups of thyrotoxic and euthyroid goiter, correlation coefficients for *I*_NADH_ *versus I*_NADPH_ and *K*_NADH_ *versus K*_NADPH_ were 0.21 and 0.04, respectively. However, the absence of a correlation can be explained by close values of the parameters (Fig. 1a).

For malignant and histologically unchanged tissues obtained from the same thyroid lobe, the significant differences were found for *K*_NADH_ (2.8-fold higher in cancer nodes, *p* = 0.008), *I*_NADPH_ (7.8-fold higher in cancer nodes, *p* = 0.012), and *K*_NADPH_ (2.8-fold higher in cancer nodes, *p* = 0.050).

The basal lucigenin-enhanced luminescence *I*_0_ was different in the subgroup of high activity of microsomal reductases (*n* = 6) compared to the control group (*p* = 0.001) (Table 2). Based on both *I*_NADH_ and *K*_NADH_, the tissue CYB5R activity in the groups of euthyroid goiter, toxic goiter, and the low-activity reductase PTC subgroup did not differ significantly (Table 1). For the high-activity reductase PTC subgroup, mean *I*_NADH_ value was 4.7-fold higher than in the control group (*p*<0.001) and 3.6-fold higher than in the thyrotoxic goiter group (*p* = 0.002). For *K*_NADN_, the mean values differed by a factor of 6.2 (*p*<0.001) and 4.9 (*p* = 0.001).

Unlike CYB5R, the CYPOR activity in thyrotoxic goiter was 7.3-fold higher than in the control group (*p* = 0.011), the activation coefficient *K*_NADPH_ was 5.8-fold higher (*p* = 0.031) (Table 1). *I*_NADPH_ values in the high-activity reductase PTC subgroup was 22.2-fold higher than in the control group (*p* = 0.001) and 3.0-fold higher than in the thyrotoxic-goiter group (*p* = 0.003). As for the activation coefficient, it was 17.8-fold (*p* = 0.001) and 3.0-fold higher (*p* = 0.009), respectively. The low-activity reductase PTC subgroup did not differ the control group and the thyrotoxic-goiter group (Table 2).

**Table 1.** The activity of cytochrome b5-reductase and cytochrome P450 reductase in malignant and histologically unchanged tissue in PTC

| Index | Histologically unchanged tissue ($n = 12$) | | Cancer tissue ($n = 12$) | | $p$ |
| --- | --- | --- | --- | --- | --- |
| | $\bar{x}$ | $S$ | $\bar{x}$ | $S$ | |
| $I_{\text{NADH}}$ | 1.37 | 0.72 | 4.32 | 3.49 | 0.128 |
| $K_{\text{NADH}} = (I_{\text{NADH}} - I_0)/I_0$ | 10.14 | 7.57 | 28.95 | 7.61 | **0.008** |
| $I_{\text{NADPH}}$ | 1.07 | 0.96 | 7.83 | 6.86 | **0.012** |
| $K_{\text{NADPH}} = (I_{\text{NADPH}} - I_0)/I_0$ | 11.01 | 9.36 | 31.55 | 17.41 | **0.050** |

**Table 2.** The activity of tissue cytochrome b5 reductase and cytochrome P450 reductase in euthyroid goiter, thyrotoxic goiter and PTC

| Index | Euthyroid goiter ($n = 10$) | | Thyrotoxic goiter ($n = 14$) | | PTC, low activity of microsomal reductases ($n = 6$) | | PTC, high activity of microsomal reductases ($n = 6$) | |
| --- | --- | --- | --- | --- | --- | --- | --- | --- |
| | $\bar{x}$ | $S$ | $\bar{x}$ | $S$ | $\bar{x}$ | $S$ | $\bar{x}$ | $S$ |
| $I_0$ | 0.11 | 0.03 | 0.15 | 0.07 | 0.13 | 0.05 | 0.24 ($p = 0.001^*$)<br>($p = 0.071^{**}$) | 0.07 |
| $I_{\text{NADH}}$ | 0.52 | 0.14 | 0.69 | 0.54 | 0.47 | 0.11 | 2.45 ($p < 0.001^*$. $p = 0.002^{**}$) | 1.43 |
| $K_{\text{NADH}} = (I_{\text{NADH}} - I_0)/I_0$ | 3.77 | 1.92 | 4.74 | 3.50 | 2.62 | 1.01 | 23.29 ($p < 0.001^*$. $p = 0.001^{**}$) | 14.38 |
| $I_{\text{NADPH}}$ | 0.44 | 0.30 | 3.22 ($p = 0.011^*$) | 0.34 | 1.09 | 0.34 | 9.78 ($p = 0.001^*$. $p = 0.003^{**}$) | 5.71 |
| $K_{\text{NADPH}} = (I_{\text{NADPH}} - I_0)/I_0$ | 2.53 | 2.02 | 14.83 ($p = 0.031^*$) | 7.71 | 7.83 | 5.36 | 45.16 ($p = 0.001^*$. $p = 0.009^{**}$) | 13.41 |

\* — significant differences related to the control group
\*\* — significant differences related to the thyrotoxic goiter group

**Table 3.** The activity of tissue cytochrome b5 reductase and cytochrome P450 reductase in Hürthle cell and follicular adenomas

| Index | Follicular adenoma ($n = 4$) | | Other cases of nontoxic goiter ($n = 6$) | | Hürthle cell adenoma ($n = 3$) | | Other cases of toxic goiter ($n = 11$) | |
| --- | --- | --- | --- | --- | --- | --- | --- | --- |
| | $\bar{x}$ | $S$ | $\bar{x}$ | $S$ | $\bar{x}$ | $S$ | $\bar{x}$ | $S$ |
| $I_0$ | 0.08 | 0.02 | 0.20 | 0.18 | 0.18 | 0.07 | 0.15 | 0.08 |
| $I_{\text{NADH}}$ | 0.38* ($p = 0.03$) | 0.04 | 0.63 | 0.09 | 1.21 | 0.94 | 0.46 | 0.31 |
| $K_{\text{NADH}} = (I_{\text{NADH}} - I_0)/I_0$ | 2.52* ($p = 0.04$) | 0.38 | 4.97 | 1.81 | 6.97 | 5.16 | 4.19 | 2.78 |
| $I_{\text{NADPH}}$ | 0.16* ($p = 0.05$) | 0.03 | 0.86 | 0.67 | 6.98 | 2.54 | 2.81 | 2.69 |
| $K_{\text{NADPH}} = (I_{\text{NADPH}} - I_0)/I_0$ | 1.11* ($p = 0.04$) | 0.66 | 4.20 | 1.55 | 33.43*<br>* ($p = 0.05$) | 12.44 | 11.65 | 10.71 |

\* — differences in the subgroup of follicular adenoma related to other cases of nontoxic goiter
\*\* — differences in the subgroup of Hürthle cell adenoma related to other cases of toxic goiter

No correlation was found between the activity of CYB5R and serum thyroxine for euthyroid goiter and thyrotoxic goiter. The correlation coefficient between the CYPOR activity and the level of thyroxine for euthyroid goiter was *r* = 0.75; for thyrotoxic goiter *r* = 0.55.

The activity of microsomal reductases for follicular and Hürthle cell adenomas were compared with other cases of nontoxic and thyrotoxic goiter (Table 2). For Hürthle cell adenoma, only *K*_NADPH_ was about 3-fold higher than for other cases of toxic goiter. For follicular adenomas, all the indices were 2-4-fold lower than for other cases of nontoxic goiter.

It was interesting to compare the reductase indices for follicular adenoma and the low-activity PTC. Significant differences were obtained for *I*_0_ (*p* = 0.05), *I*_NADPH_ (*p* = 0.03) and *K*_NADPH_ (*p* = 0.05), i.e. for CYPOR, but not for CYB5R activity.

## Discussion

The key results of the study are as follows: (1) follicular adenoma is characterized by 2–4 times lower activity of microsomal reductases compared to other cases of nontoxic goiter; (2) CYPOR activity is significantly increased in thyrotoxic nodes compared to the nontoxic goiter group; (3) in papillary thyroid cancer, the activity of CYB5R and CYPOR in malignant tissue is significantly higher than in morphologically unchanged tissues nearby, and the activities of CYB5R and CYPOR strongly correlate with each other; (4) in PCT, there are subgroups with low and high activity of microsomal reductases.

It is difficult to consider all aspects of the obtained results. Here, we will focus on ROS-homeostasis and oxidative stress since microsomal respiratory chains are the sources of ROS and maintains the intracellular oxidative-antioxidant and redox balance.

ROS homeostasis and redox balance are of importance for thyroid cells, since the synthesis of thyroid hormones requires hydrogen peroxide — thyroid peroxidase oxidizes inorganic iodide to iodinium ion (I^+^) and hypoiodous acid (HIO), which then attach to thyroglobulin. H_2_O_2_ is generated by special double NADPH oxidases (DUOX1 and DUOX2) located on the apical membrane of thyrocytes. DUOX catalyzes two-stage synthesis — the superoxide radical anion turns immediately into hydrogen peroxide [16]. The regulation mechanisms for these enzymes have not been fully studied, but TSH participates in this process [17].

Besides DUOX1/2, NADPH oxidase 4 (NOX4) plays an important role as a source of superoxide. NOX4 does not require a stimulus and produces ROS constantly. In addition to the superoxide anion radical, small amounts of hydrogen peroxide are formed [18]. NOX4 is located on the cytoplasmic membrane and on the membranes of the endoplasmic reticulum, mitochondria, and nuclei, which determines its multilateral effect [19]. NOX4 activity decreases with increasing concentration of hydrogen peroxide due to the oxidation of cysteine thiol groups, which can be a regulating mechanism in oxidative stress and hypoxia [20]. Presumably, this is NOX4 and its ROS that play the most important role in carcinogenesis [21]. Increased transcription of NOX4 and consequently increased hydrogen peroxide result are characteristic for various types of cancer, including thyroid cancer [21] [22].

Cytochrome b5 reductase (CYB5R) exists in two isoforms. This enzyme reduces cytochrome b5, the endpoint of the NADH-dependent microsomal respiratory chain. In red blood cells, the soluble isoform of cytochrome b5 reductase (methemoglobin reductase) is involved in the reduction of methemoglobin to hemoglobin. The microsomal isoform is located in the cytoplasmic membrane, membranes of the endoplasmic reticulum and mitochondria, Golgi apparatus, peroxisomes, nuclei, sarcoplasmic reticulum and neuronal synapses. NADH-dependent microsomal chain involves in oxidation of xenobiotics and carcinogens, biodegradation of antitumor drugs, elongation of fatty acids, cholesterol anabolism, redox signaling in neurons [8]. When cytochrome b5 is absent, CYB5R transfers electrons to other acceptors, maintaining reduced coenzyme Q10 and ascorbate [23]. With ascorbate, the NADH-dependent cytochrome b5 reductase system protects thyroid hormones from oxidative degradation [24]; reduced coenzyme Q10 prevents lipid peroxidation in plasma membranes and apoptosis [23]. Thus, the NADH-dependent cytochrome b5 reductase system can be considered as a part of the intracellular antioxidant system.

On the other hand, when NADH-dependent microsomal chain oxidizes xenobiotics, ROS are formed resulting in lipid peroxidation and DNA damage [25] [26]. The increased CYB5R activity in the plasma membrane leads to excess production of the superoxide anion radical, which induces the irreversible stages of apoptosis of cerebellar granular neurons [27]. The expression of CYB5R in HeLa cells is inhibited by H_2_O_2_, but then increased via the unknown mechanisms [28].

The participation of CYB5R in thyroid hormonogenesis is studied in a few works. A case of nontoxic goiter (TSH = 17.8 μmol/L) was described, where the activity of microsomal cytochrome b5 reductase had been significantly reduced in thyroid tissue homogenates due to an impaired biosynthesis of FAD (which is a coenzyme of CYB5R) from riboflavin. Therefore, organification of iodine was decreased [29]. In our study, there were no patients with hypothyroidism, and we could not to prove a significant increase in the CYB5R activity in thyrotoxicosis (*p* = 0.071). It is likely that an increase in CYB5R activity may by compensatory in Graves’ disease.

The information on the role of CYB5R in carcinogenesis is extremely poor and contradictory. The study [30] [31] demonstrate that CYB5R2 has an oncosuppressive effect due to the suppression of neoangiogenesis. Contrary data was obtained by the authors of [10], who proved the key role of CYB5R3 in extravasation and colonization of cancer cells after intravenous administration to mice. Knockdown of the CYB5R3 gene in cancer cells led to a significant decrease in tumor mass in the lungs of mice. The authors explained the cellular effects of knockdown of CYB5R3 by the relationship with signaling TGF-β-dependent pathways, HIF-α-dependent pathways, and apoptosis. The increased CYB5R3 in biopsy specimens correlated with a short relapse-free interval and survival of patients with estrogen receptor negative cancers.

Cytochrome P450 reductase transfers electrons from NADPH to cytochrome P450. Other substances also can be electron acceptors such as quinones [32], cytochrome b5, heme oxygenase, and other enzymes [9]. The NADPH-dependent cytochrome P450 system is considered the main intracellular source of ROS (superoxide radical anion and hydrogen peroxide) along with the mitochondrial respiratory chain. Defects in the cytochrome P450-dependent chain lead to a marked ROS disbalance, lipid peroxidation and apoptosis. Therefore, functioning of this chain is strongly controlled by gene expression, protein interactions, and oxidative stress by the feedback mechanism [33]. ROS produced by the NADPH-dependent cytochrome P450 system play a significant role in carcinogenesis [34]. An increased CYPOR expression results in the increased ROS production [35].

Here, we have observed mor marked effects for CYPOR rather than CYB5R. We considered patients with nontoxic goiter as a control group. However, for follicular adenomas, a decrease in the activity of both reductases was shown. Interestingly, the CYPOR activity in follicular adenoma was significantly lower compared even to the PTC group with low reductase activity. In thyrotoxicosis, CYPOR activity was significantly increased. This corresponds to the fact that T3 activates CYPOR transcription [36] [37] [38]. It can be assumed that increased hydrogen peroxide activates DUOX and hormonogenesis [39].

In the PTC group, half of the cases were characterized by high CYB5R3 activity. This is in the line with the study [10], where increased CYB5R3 activity correlated with poor survival prognosis and metastasis. The reasons for the increased CYB5R3 activity of are not clear, but we can propose two reasons. The CYB5R3 activity can be increased to compensate oxidative stress caused, for example, by NOX4. On the other hand, the CYB5R3/CYB5 microsomal chain can be an extra source of ROS, which can aggravate the development of cancer.

In PTC, we have found more marked effects for CYPOR rather than CYB5R. Perhaps, this is due to the more significant role of the cytochrome P450 system as a generator of reactive oxygen species. To answer how the increase in the activity of microsomal reductases is related to ROS homeostasis can be given by a special study with simultaneous measurement of NADH/NADPH-stimulated chemiluminescence, expression of the CYB5R/CYPOR gene and visualization of intracellular ROS.

## Conclusion

To sum, the activity of CYB5R and CYPOR increases several times in thyrotoxicosis, increases significantly in some cases of papillary thyroid cancer, and reduces in follicular adenoma. The changes in CYPOR activity are manifested more clearly. In future, we intend to compare the microsomal activity with long-term outcomes, recurrence, and metastasis. The data obtained are of importance for fundamental and clinical oncology.

### Statement of Ethics

All the participants of the study have given their written informed consent. The study was approved by the Ethics Committee of Inozemtsev City Clinical Hospital (Approval No. 12, 11.10.2019).

## Data Availability

The data used to support the findings of this study can be requested from the corresponding author.

## Contribution

Conceptualization and design — E.V. Proskurnina, I.V. Panteleev; methodology, E.V. Proskurnina, clinical data — I.V. Panteleev, E.V. Svetlov; resources — A.E. Mitichkin, investigation — M.V. Fedorova, M.M. Sozarukova; interpretation and original draft preparation — E.V. Proskurnina; review and editing — I.V. Panteleev. All authors have read and agreed to the published version of the manuscript.

